# Ancestry-specific regulatory and disease architectures are likely due to cell-type-specific gene-by-environment interactions

**DOI:** 10.1101/2023.10.20.23297214

**Authors:** Juehan Wang, Steven Gazal

## Abstract

Multi-ancestry genome-wide association studies (GWAS) have highlighted the existence of variants with ancestry-specific effect sizes. Understanding where and why these ancestry-specific effects occur is fundamental to understanding the genetic basis of human diseases and complex traits. Here, we characterized genes differentially expressed across ancestries (ancDE genes) at the cell-type level by leveraging single-cell RNA-seq data in peripheral blood mononuclear cells for 21 individuals with East Asian (EAS) ancestry and 23 individuals with European (EUR) ancestry (172K cells); then, we tested if variants surrounding those genes were enriched in disease variants with ancestry-specific effect sizes by leveraging ancestry-matched GWAS of 31 diseases and complex traits (average *N* = 90K and 267K in EAS and EUR, respectively). We observed that ancDE genes tend to be cell-type-specific, to be enriched in genes interacting with the environment, and in variants with ancestry-specific disease effect sizes, suggesting the impact of shared cell-type-specific gene-by-environment (GxE) interactions between regulatory and disease architectures. Finally, we illustrated how GxE interactions might have led to ancestry-specific *MCL1* expression in B cells, and ancestry-specific allele effect sizes in lymphocyte count GWAS for variants surrounding *MCL1*. Our results imply that large single-cell and GWAS datasets in diverse populations are required to improve our understanding on the effect of genetic variants on human diseases.

## Introduction

Multi-ancestry genome-wide association studies (GWAS) have highlighted that, despite the strong correlation of causal effect sizes across ancestries ^1–9^, a non-negligible fraction of causal variants have ancestry-specific effect sizes, likely due to gene-by-environment (GxE) interactions ^7–9^. Knowing where and why ancestry-specific effects of disease risk variants occur is fundamental for understanding the genetic basis of human diseases and for improving the portability of polygenic risk scores across ancestries ^6^.

Differences in gene regulation across ancestries have been observed at different regulatory levels (such as gene expression ^10–18^, eQTL effect sizes ^19–21^, methylation ^22,23^ and enhancer activity ^24^) and could inform which variants have ancestry-specific disease effect sizes. Indeed, gene regulation differences can also be due to GxE (e.g., ancestry-specific eQTL effect sizes), and variants with ancestry-specific disease effect sizes tend to be enriched in regulatory regions and around genes differentially expressed in tissues interacting with the environment ^7^. However, investigating if ancestry-specific regulatory and disease architectures are correlated (because of shared GxE mechanisms) has been challenging for multiple reasons. First, there is a limited availability of ancestry matched GWAS and functional datasets from non-European descent. Second, while regulatory differences (such as ancestry-specific gene expression) could be due to GxE, they can also be the consequences of differences in allele frequencies due to genetic drift, different answers to the ancestry environment with no genetic mediation, or batch effects due to how multi-ancestry data have been collected ^12^. Finally, as gene regulation tend to be cell-type-specific ^25,26^, it is unclear which cell types are the most subject to ancestry-specific gene regulation and disease effect sizes.

Here, we aim to characterize genes differentially expressed across ancestries (ancDE genes) at the cell-type level, and to test if ancDE genes are enriched in disease variants with ancestry-specific effect sizes. We leveraged single-cell RNA-seq (scRNA-seq) data in peripheral blood mononuclear cells (PBMCs) for 21 individuals with East Asian (EAS) ancestry and 23 with European (EUR) ancestry ^27^ (172,385 cells analyzed across 7 main cell types), and ancestry-matched GWAS of 31 diseases and complex traits ^7^ (average *N* = 90K and 267K in EAS and EUR, respectively). We observed that ancDE genes tend to be cell-type-specific, to be enriched in genes interacting with the environment and in variants with ancestry-specific disease effect sizes, suggesting the impact of shared cell-type-specific GxE interactions between regulatory and disease architectures. Our results imply that large single-cell and GWAS datasets in diverse populations are required to improve our understanding of human diseases.

## Results

### Overview of the methods

We characterized ancDE genes by leveraging scRNA-seq data in PBMCs from 21 and 23 healthy females of EAS and EUR ancestry, respectively ^27^. We focused on the 7 most abundant cell types, with number of cells varying between 2,284 and 71,207 across them (172,385 total cells across the 7 cell types): B cells (B), Natural killer cells (NK), CD4+ and CD8+ T cells (T4 and T8, respectively), conventional dendritic cells (cDC) and classical and non-classical monocytes (cM and ncM, respectively) (**Figure 1A**). Cell-type proportions were not significantly different across the two populations (**Figure 1B** and **Supplementary Table 1**). Differential expression across ancestries was tested independently in each cell type for each expressed gene using a Poisson linear mixed-effects model with the donor as a random effect, and ancestry and multiple covariates as fixed effects.

**Figure 1:**
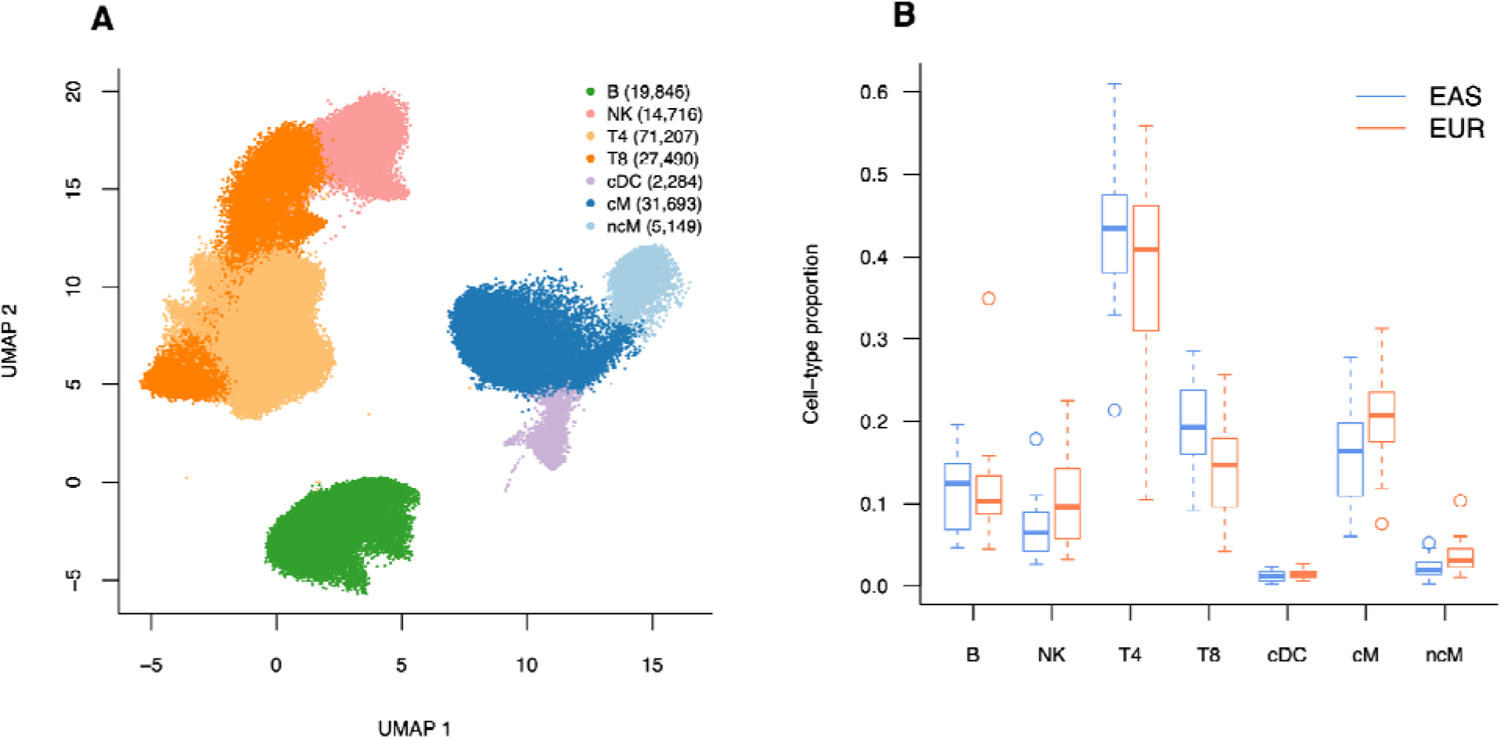
An immune multi-ancestry single-cell dataset. **(A)** We report UMAP coordinates and assignment of 172,385 cells to 7 immune cell types ^27^: B cells (B), Natural killer cells (NK), CD4+ and CD8+ T cells (T4 and T8, respectively), conventional dendritic cells (cDC) and classical and non-classical monocytes (cM and ncM, respectively). The number of cells in each cell type is reported in the legend. **(B)** We report cell-type proportions across 21 EAS and 23 EUR individuals; we did not observe significant differences (*P* < 0.05/7) of cell-type proportions across ancestries (min *P* = 0.03 in cM; **Supplementary Table 1**). The median value of each proportion is displayed as a band inside each box. Boxes denote values in the second and third quartiles. The length of each whisker is 1.5 times the interquartile range (defined as the height of each box). All values lying outside the whiskers are considered to be outliers.

We tested if SNPs surrounding ancDE genes (i.e., SNPs in a +/- 100kb window) were enriched in ancestry-specific effect sizes by using the S-LDXR method ^7^. For a given annotation *C*, S-LDXR reports the enrichment of squared multi-ancestry genetic correlation (λ^2^), which is defined as

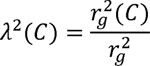

where *r_g_^2^* is the squared correlation of the per-allele effect sizes in the two populations (i.e., squared multi-ancestry genetic correlation), and *r_g_^2^(C)* is the squared multi-ancestry genetic correlation computed within SNPs in *C*. We note that λ^2^ is not impacted by allele frequency differences across the two populations (as it computes correlation of per-allele effect sizes and not effect sizes on normalized genotypes), and that S-LDXR estimates of λ^2^ were unbiased for annotations with allele frequency differences in simulations (and conservative for other annotations) ^7^. Here, we applied S-LDXR with the annotations of the baseline-LD-X model on 31 diseases and complex traits ^7^ (average *N* = 90K and 267K in EAS and EUR, respectively); results were meta-analyzed across 20 approximately independent traits, including 10 approximately independent blood and immune-related traits (**Supplementary Table 2**). We constructed SNP-annotations based on gene sets by selecting all SNPs falling in 100-kb windows on either side of the gene bodies.

Further details are provided in **Methods**. We have released differential gene expression results and corresponding S-LDXR annotations, as well as code to replicate our analyses (Data and code availability).

### Cell-type-specificity of genes differentially expressed between East-Asian and European populations

We tested differential gene expression in 7 immune cell types within 21 and 23 healthy individuals of EAS and EUR ancestry, respectively. We detected between 8 and 160 ancDE genes with false discovery rate (FDR) < 0.05 (396 ancDE genes across the 7 cell types, 320 unique genes; **Supplementary** Fig. 1 and **Supplementary Table 3**). As the number of significant genes per cell types correlated with the number of tested cells (*r* = 0.60), we restricted further analyses to the 100 genes with the smallest *P* values within each cell type, leading to a list of 545 unique ancDE genes (including only 15 genes of the major histocompatibility complex (MHC) region) (**Supplementary Table 4**). All further analyses were replicated using the 200 and 500 genes with the smallest *P* values within each cell type.

Out of the 545 unique ancDE genes, 452 (83%) were differentially expressed in a single cell type, suggesting high cell-type-specificity of differential expression across ancestries (**Figure 2A**). Within each cell type, between 53% and 72% of ancDE genes were specific to this cell type (**Figure 2B**). Genes differentially expressed in at least two cell types tend to cluster between lymphoid (NK, B, T4 and T8) and myeloid cell types (cM, ncM, and cDC) (e.g., 28 genes are differentially expressed in both T4 and T8 cell types, and 18 genes were differentially expressed in cM and nCM cells). Within the ancDE genes differentially expressed in at least 2 cell types, a large fraction (86 out of 93; 92%) had consistent directions across the cell types (i.e., overexpressed or underexpressed in all the cell types) (**Supplementary Table 5**).

**Figure 2:**
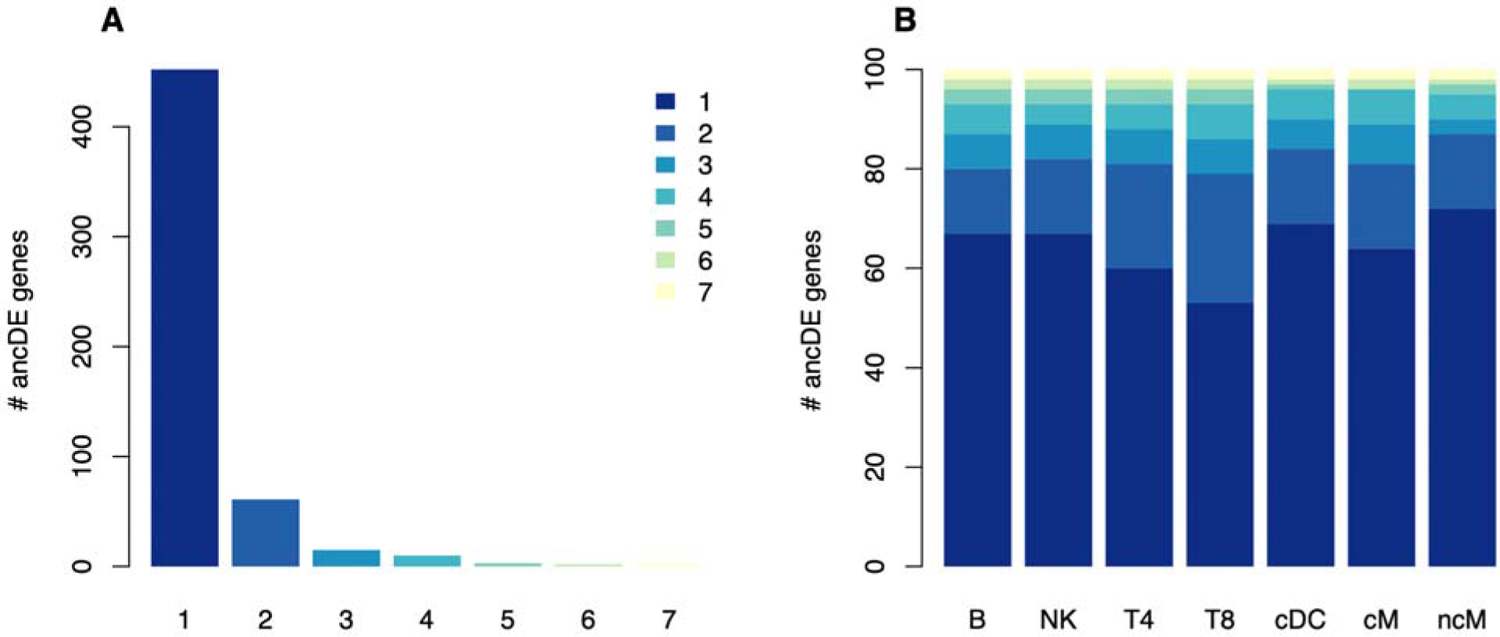
Cell-type-specificity of ancDE genes. (**A**) We report the number of cell-type-specific ancDE genes (top 100 smallest *P* values for each cell type) shared across all the cell types. We observed that 83% of ancDE genes were differentially expressed in a single cell type. (**B**) For each cell type, we report the number of ancDE genes shared across all the cell types. List of ancDE genes is reported in **Supplementary Table 4**. Across the cell types, between 53% and 72% of their ancDE genes were cell-type-specific. Similar patterns were observed when defining ancDE genes using the FDR 5% threshold, and the 200 and 500 smallest *P* values (**Supplementary** Figs. 1-3) and for genes differentially expressed in males and females ^28^ (**Supplementary** Fig. 4).

To validate that the cell-type-specificity of ancDE genes was not an artifact of a relatively low number of samples (despite the high number of cells), we performed the following supplementary analyses. First, we replicated this observation when defining ancDE genes using the FDR 5% threshold, and the 200 and 500 smallest *P* values: 85%, 82%, and 77% of unique ancDE genes were differentially expressed in a single cell type, respectively (**Supplementary** Figs. 1-3). Second, we leveraged a larger scRNA-seq with 416 EUR males and 565 EUR females ^28^ (1,175,543 cells across 7 similar cell types) and identified the top 100 genes the most significantly differentially expressed in both sex for each cell type (578 unique sexDE genes) (see **Methods**). We observed that 86% of sexDE genes were differentially expressed in a single cell type, confirming that environment differences impact gene expression at the cell-type-specific level (**Supplementary** Fig. 4 and **Supplementary Table 6**).

### AncDE genes are enriched in genes interacting with the environment

We next sought to investigate if ancDE genes tend to be driven by environmental differences (i.e., GxE interactions of their eQTLs, or different answers to environments with no genetic mediation), genetic differences (population differences in allele frequencies of their eQTLs), or both.

First, by performing gene ontology enrichment analyses ^29^, we observed that the 545 ancDE genes were enriched in genes involved in immune response to the environment (FDR corrected *P* of 7.88 × 10^-4^ for *leukocyte activation* (GO:0045321) pathway; **Supplementary Table 7**). At the cell-type level, we found the most significant enriched GO categories in ncM cells (e.g., FDR corrected *P* of 7.01 × 10^-7^ *myeloid cell activation involved in immune response* (GO:0002275) pathway), NK cells (FDR corrected *P* of 1.7 × 10^-5^ for *interferon-gamma-mediated signaling* (GO:0060333) pathway) and cM cells (FDR corrected *P* of 1.48 × 10^-2^ for *neutrophil activation involved in immune response* (GO:0002283) pathway) (**Supplementary Table 7**). We detected even more significantly enriched pathways involved in immune response when defining ancDE genes by using the 200 and 500 smallest *P* values (such as FDR corrected *P* of 4.50 × 10^-5^ and 2.22 × 10^-6^ for *response to external biotic stimulus* (GO:0043207) and *viral process* (GO: 0016032) pathways, respectively). Similar conclusions were obtained when removing MHC genes from the analysis (**Supplementary Table 7**).

Second, we investigated if ancDE genes were due to allele frequency differences of their eQTLs by leveraging cell-type-specific eQTLs from 982 EUR individuals ^28^; we note that our dataset was underpowered to detect ancestry-specific eQTLs and that there is (to our knowledge) no existing large EAS PBMC single-cell dataset available (see Discussion). We determined that nearly half of cell-type-specific ancDE genes (47%; 329 out of 700) have at least one independent EUR eQTLs in the corresponding cell type, which is 3.1 times more than what we would expect by chance (**Fig 3A**). Interestingly, these eQTLs tend to have extremely high fixation index (*F_st_*) across EAS and EUR reference populations ^30^ (mean *F_st_* = 0.21 across all ancDE gene eQTLs vs. mean *F_st_*= 0.10 across eQTLs of all expressed genes, *P* = 1 × 10^-28^ for difference; **Fig 3B**). When replicating differentially expression analyses for ancDE genes for which genotypes of the eQTLs were available (273 out of 329), we found that 71% of these genes (194 out of 273) do not remain in the top 100 most significantly differentially expressed genes after conditioning on their eQTLs (**Fig 3C**). These results suggest that at least a third of ancDE genes (0.47 x 0.71) are driven by allele frequency differences of their eQTLs across ancestries. We replicated all our analyses and conclusion by defining ancDE genes using an FDR 5% threshold, as well as the 200 and 500 genes with the smallest differentially gene expression *P* values within each cell type (**Supplementary** Figs. 5-7).

**Figure 3:**
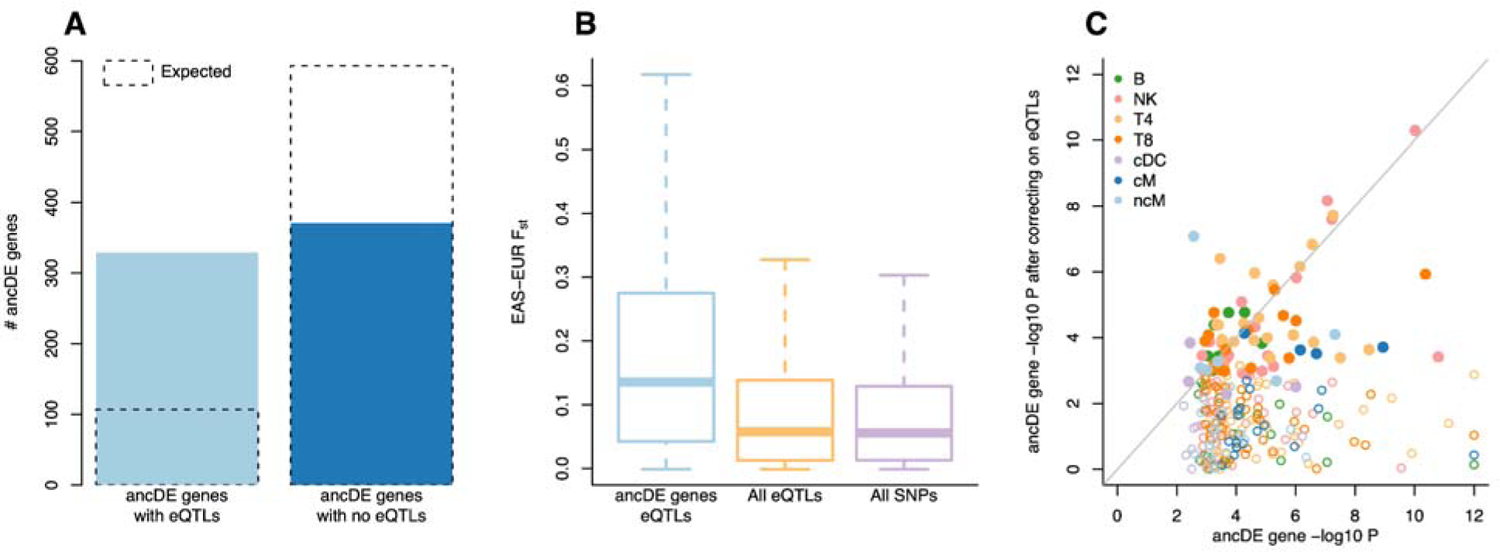
AncDE genes are driven by allele frequency differences of their eQTLs. (**A**) We report the number of cell-type-specific ancDE genes with at least one EUR eQTLs and without eQTLs in the corresponding cell type. Dotted boxes represent the number of ancDE genes that would have been observed by chance. (**B**) We report mean fixation index (*F_st_*) across EAS and EUR reference populations ^30^ for all ancDE gene eQTLs, eQTLs of all expressed genes, and all SNPs. (**C**) Scatter plot of ancDE genes -log_10_(*P*) before and after conditioning on their eQTLs. Solid points represent ancDE genes that remain in the top 100 most significantly differentially expressed genes after conditioning on their eQTLs.

Finally, we investigated if allele frequency differences of eQTLs across ancestries were more likely due to adaptation to new environments or to genetic drift. We observed that ancDE genes with eQTLs were also enriched in genes involved in immune response (minimum FDR corrected *P* of 1.31 × 10^-2^, 1.68 × 10^-2^, 4.13 × 10^-4^ and 1.43 × 10^-5^ for *myeloid cell activation involved in immune response* (GO:0002275), *peptide antigen binding* (GO:0042605) pathway, *response to interferon-gamma* (GO:0034341) and *peptide antigen binding* (GO:0042609) pathways when considering the FDR 5% threshold, and the 100, 200 and 500 most significant ancDE genes, respectively; **Supplementary Table 7**), suggesting that allele frequency differences of their eQTLs might have been driven by adaptation to new environments rather than genetic drift. AncDE genes without eQTLs were enriched in genes involved in immune response but significance did not pass the FDR 5% threshold (except when considering the top 500 most significant ancDE genes; **Supplementary Table 7**). Finally, we observed that 27% of cell-type-specific sexDE genes (186 out of 700) have at least one independent eQTLs (**Supplementary** Fig. 8), demonstrating that genes with eQTLs can be differentially expressed even without differences in allele frequencies.

All together, these results suggest that ancDE genes are enriched in genes interacting with the environment. At least a third of ancDE genes could be due to allele frequency differences of their eQTLs, although it is very likely that a large fraction of these eQTLs is enriched in GxE interactions.

### AncDE genes are enriched in ancestry-specific causal effect sizes of complex traits

We created a SNP-annotation for the 545 unique ancDE genes (annotation representing 4.8% of investigated common SNPs) and analyzed it using S-LDXR on 31 diseases and complex traits and meta-analyzed results across 20 approximately independent traits. We observed that SNPs in ancDE genes were significantly enriched in SNP-heritability (*h*^2^) in both populations (*h*^2^ enrichment = 2.07 ± 0.18, *P* = 3 × 10^-9^ in EAS; *h*^2^ enrichment = 1.71 ± 0.13, *P* = 2 × 10^-8^ in EUR) highlighting the impact of ancDE genes on human diseases and complex traits.

We determined that SNPs within ancDE genes were extremely depleted of squared multi-ancestry genetic correlation (λ^2^ = 0.69 ± 0.04, *P* = 6 × 10^-13^; **Figure 4** and **Supplementary Tables 8 and 9**) and is more depleted (and most significantly depleted) than any other annotation from the baseline-LD-X model (**Supplementary Table 10**). We detected significant depletions (*P* < 0.05/31) for 6 traits, including: hematocrit (λ^2^ = 0.35 ± 0.12, *P* = 5 × 10^-8^), lymphocyte count (λ^2^ = 0.52 ± 0.12, *P* = 2 × 10^-5^) and height (λ^2^ = 0.71 ± 0.09, *P* = 5 × 10^-4^) (**Supplementary Table 8**). AncDE genes λ^2^ was also significantly lower than λ^2^ estimated on all genes (λ^2^ = 0.95 ± 0.01, *P* = 3 × 10^-9^ for difference with ancDE genes) and on genes expressed in the 7 cell types (λ^2^ = 0.91 ± 0.01, *P* = 1 × 10^-6^ for difference with ancDE genes); by performing 100 random sampling of 545 genes, we also validated that ancDE genes were significantly depleted in multi-ancestry genetic correlation (λ^2^ = 0.84 ± 0.01, *P* < 1/100 for difference with ancDE genes; **Supplementary** Fig. 9). The net contribution of the ancDE gene annotation to the covariance of effect sizes (θ, see **Methods**) was only slightly significant (*P* = 0.02; **Supplementary Table 9**), meaning that most of the multi-ancestry genetic correlation depletion of this annotation was already captured by existing annotations of the baseline-LD-X model. Indeed, we observed that the variants within the ancDE gene annotation tend to have higher values for annotations with depleted λ^2^ that are related to regulatory regions and/or selection; for example, they tend to be enriched in promoters, exons and UTRs, and to have higher background selection statistics ^31,32^ and lower predicted allele age ^32,33^ than other SNPs (**Supplementary Table 11**).

**Figure 4:**
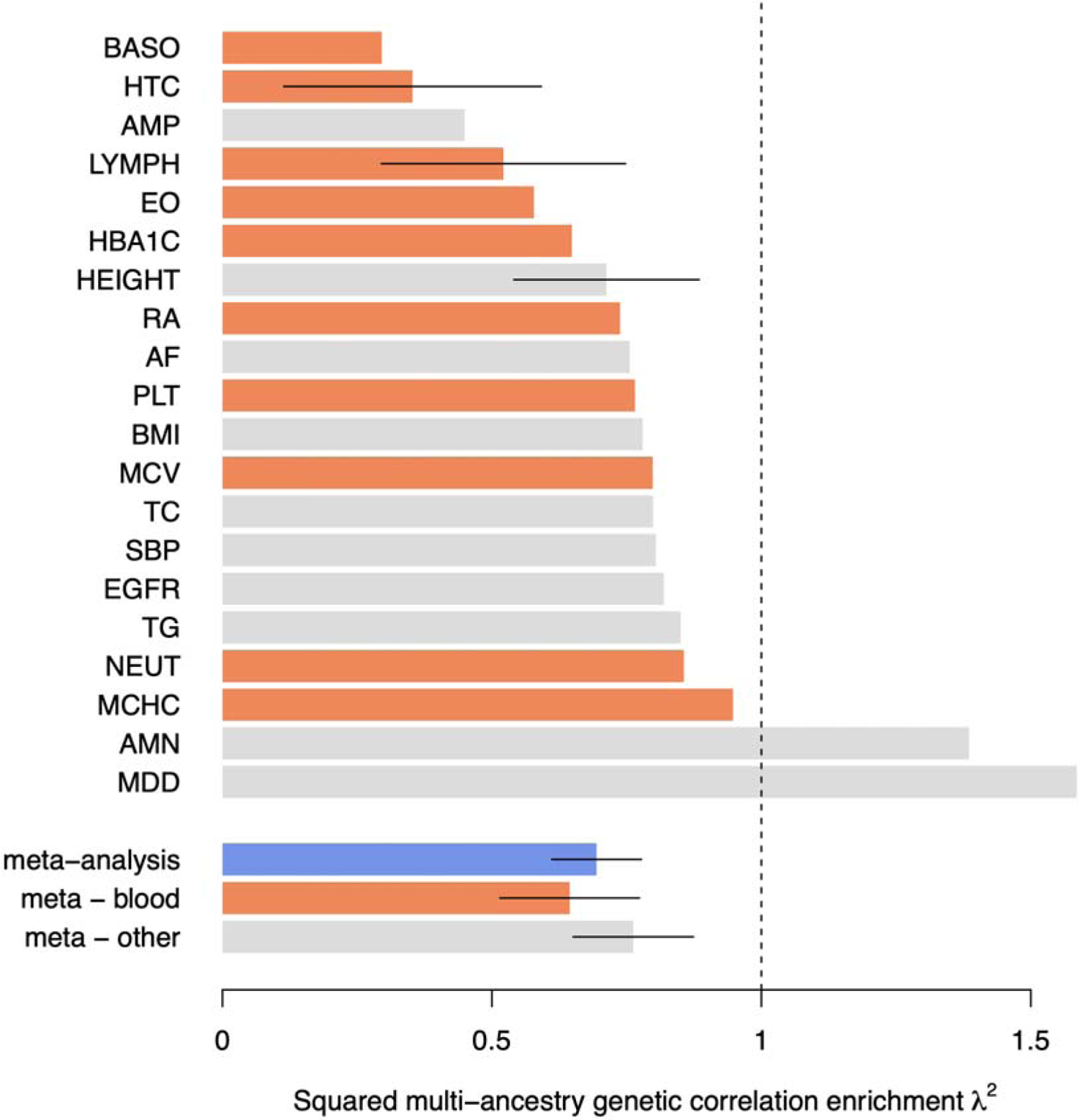
Squared multi-ancestry genetic correlation enrichment for variants surrounding ancDE genes. We report squared multi-ancestry genetic correlation enrichment (λ^2^) for each independent trait and meta-analyses results across groups of traits. Orange bars represent blood and immune-related traits, gray bars represent other traits, and the blue bar represents the meta-analysis across all traits. Error bars represent 95% confidence interval (CI) for traits with λ^2^ significantly lower than 1. Numerical results are reported in **Supplementary Tables 8-9**. AF: Atrial Fibrillation; AMN: Age at Menarche; AMP: Age at Menopause; BASO: Basophil Count; BMI: Body Mass Index; EGFR: Estimated Glomerular Filtration Rate; EO: Eosinophil Count; HBA1C: Hemoglobin A1c; HTC: Hematocrit; LYMPH: Lymphocyte Count; MCHC: MCH Concentration; MCV: Mean Corpuscular Volume; MDD: Major Depressive Disorder; NEUT: Neutrophil Count; PLT: Platelet Count; RA: Rheumatoid Arthritis; SBP: Systolic Blood Pressure; TC: Total Cholesterol; TG: Triglyceride.

As expected, λ^2^ was even smaller when meta-analyzed across 10 approximately independent blood and immune-related traits (λ^2^ = 0.64 ± 0.07, *P* = 7 × 10^-8^), and remained significantly depleted in the 10 remaining traits (λ^2^ = 0.76 ± 0.06, *P* = 3 × 10^-5^). Similar conclusions were obtained when defining ancDE genes using the FDR 5% threshold, and the 200 and 500 smallest *P* values (**Supplementary Table 9**). We rerun S-LDXR on two distinct annotations corresponding to ancDE genes with and without eQTLs, respectively (each annotation represents 2.4% and 2.5% of investigated SNPs, respectively). We observed similar depletion of squared multi-ancestry genetic correlation for the two annotations (λ^2^ = 0.65 ± 0.07 and λ^2^ = 0.66 ± 0.04, respectively) (**Supplementary Table 9**), suggesting that even if genes were differentially expressed due to allele frequency differences of their eQTLs, these genes are likely enriched in ancestry-specific causal effect sizes.

To support that genes with varying levels of expression in different environments are enriched in context-specific causal effect sizes, we extended S-LDXR to stratify squared genetic correlation between male and female GWASs and applied it to sexDE genes annotations on 17 independent traits previously identified with sex genetic correlation significantly less than 1 (ref. ^34^) (see **Methods**). We observed significant depletion of squared sex genetic correlation within our male and female GWASs (λ^2^ = 0.91 ± 0.02, *P* < 2 × 10^-7^), and similar depletion when stratifying sexDE genes with and without eQTLs (λ^2^ = 0.89 ± 0.03 and λ^2^ = 0.90 ± 0.02, respectively) (**Supplementary Table 12**).

Finally, to refine ancDE gene λ^2^ signal, we performed S-LDXR analyses by creating SNP-annotations for each of the 7 main cell types (each annotation represents between 0.8% and 1.0% of investigated SNPs). The 7 SNP-annotations were all depleted of squared multi-ancestry genetic correlation (λ^2^ < 0.78; **Figure 5** and **Supplementary Table 9**) and all cell types except T8 were significantly depleted. We observed the smallest and most significant depletion for the ancDE genes in B cells (λ^2^ = 0.35 ± 0.06, *P* = 1 × 10^-24^) and cDC cells (λ^2^ = 0.36 ± 0.10, *P* = 6 × 10^-10^). Smaller λ^2^ for the B and cDC annotations were observed when meta-analyzed across 10 approximately independent blood and immune-related traits (λ^2^ = 0.30 ± 0.07, *P* = 4 × 10^-21^ and λ^2^ = 0.21 ± 0.12, *P* = 1 × 10^-10^, respectively); this trend was not observed across the 10 remaining traits (**Supplementary** Fig. 10 and **Supplementary Table 9**). Similar conclusions were obtained when defining ancDE genes using the 200 and 500 smallest *P* values (**Supplementary** Figs. 11-12 and **Supplementary Table 9**).

**Figure 5:**
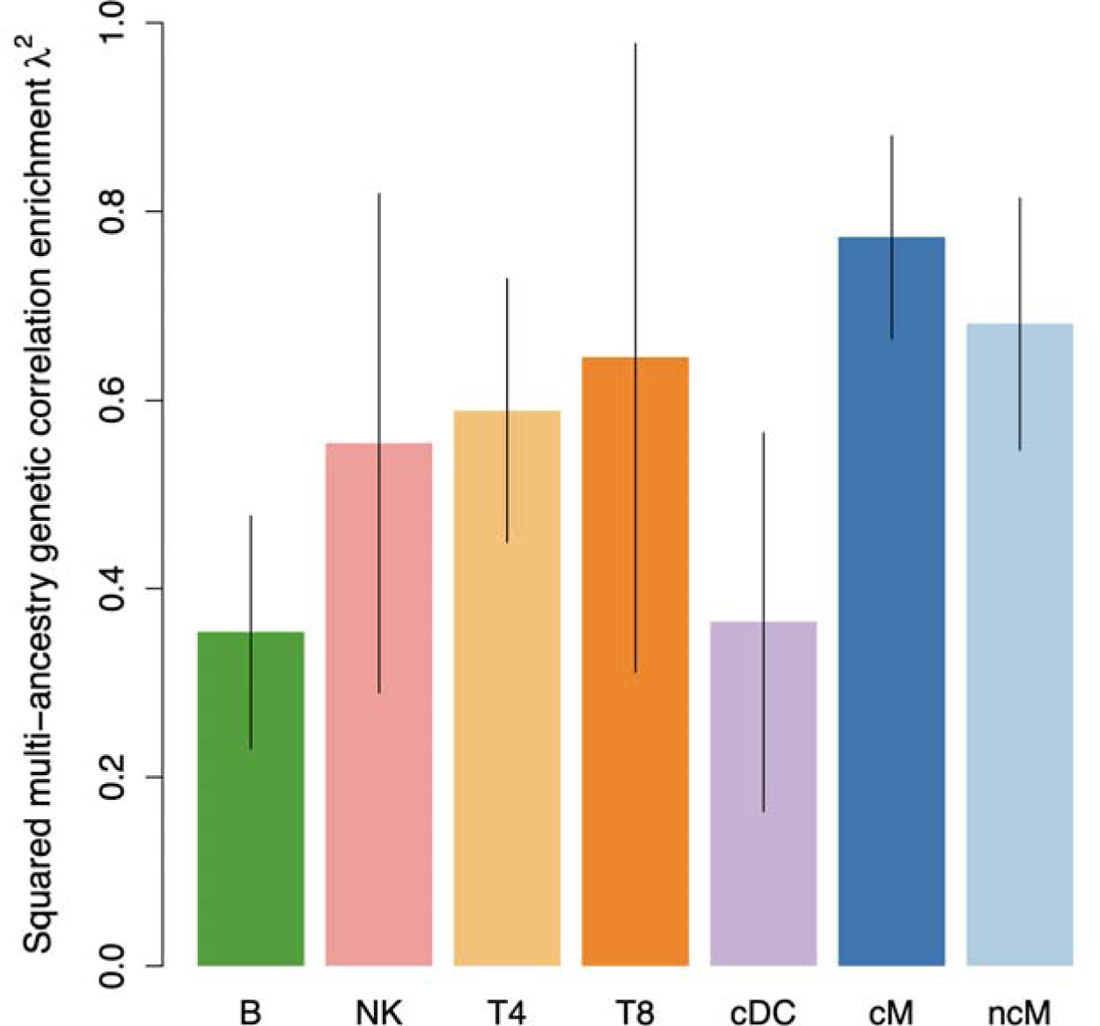
Squared multi-ancestry genetic correlation enrichment for variants surrounding cell-type-specific ancDE genes. We report squared multi-ancestry genetic correlation enrichment (λ^2^) for cell-type-specific ancDE gene annotations meta-analyzed across 20 independent traits. Error bars represent 95% CI. Numerical results are reported in **Supplementary Table 9**.

Altogether, these results demonstrate discordant causal effect sizes between EAS and EUR GWAS for variants surrounding ancDE genes, likely due to GxE interactions. The magnitude of λ^2^ was similar for genes with and without eQTLs, suggesting that even if a gene is differentially expressed because of different allele frequencies of its eQTLs, this gene is likely to also be enriched in GxE effects (as difference of allele frequencies might have been driven by adaptation). Finally, for blood and immune-related traits, we observed stronger discordant effect sizes for SNPs within ancDE genes in B cells and cDC cells, two cell types that initiate and shape the adaptive immune response to new environments.

### Illustrating ancDE genes with strong GWAS discordant effect sizes

Here, we illustrate how GxE interactions may have led to differential expression of the ancDE gene *MCL1* in B cells, different *MCL1* eQTL effect sizes in blood, and different allele effect sizes around *MCL1* in lymphocyte count (LYMPH) EAS and EUR GWASs (**Figure 6** and **Supplementary** Fig. 13).

**Figure 6:**
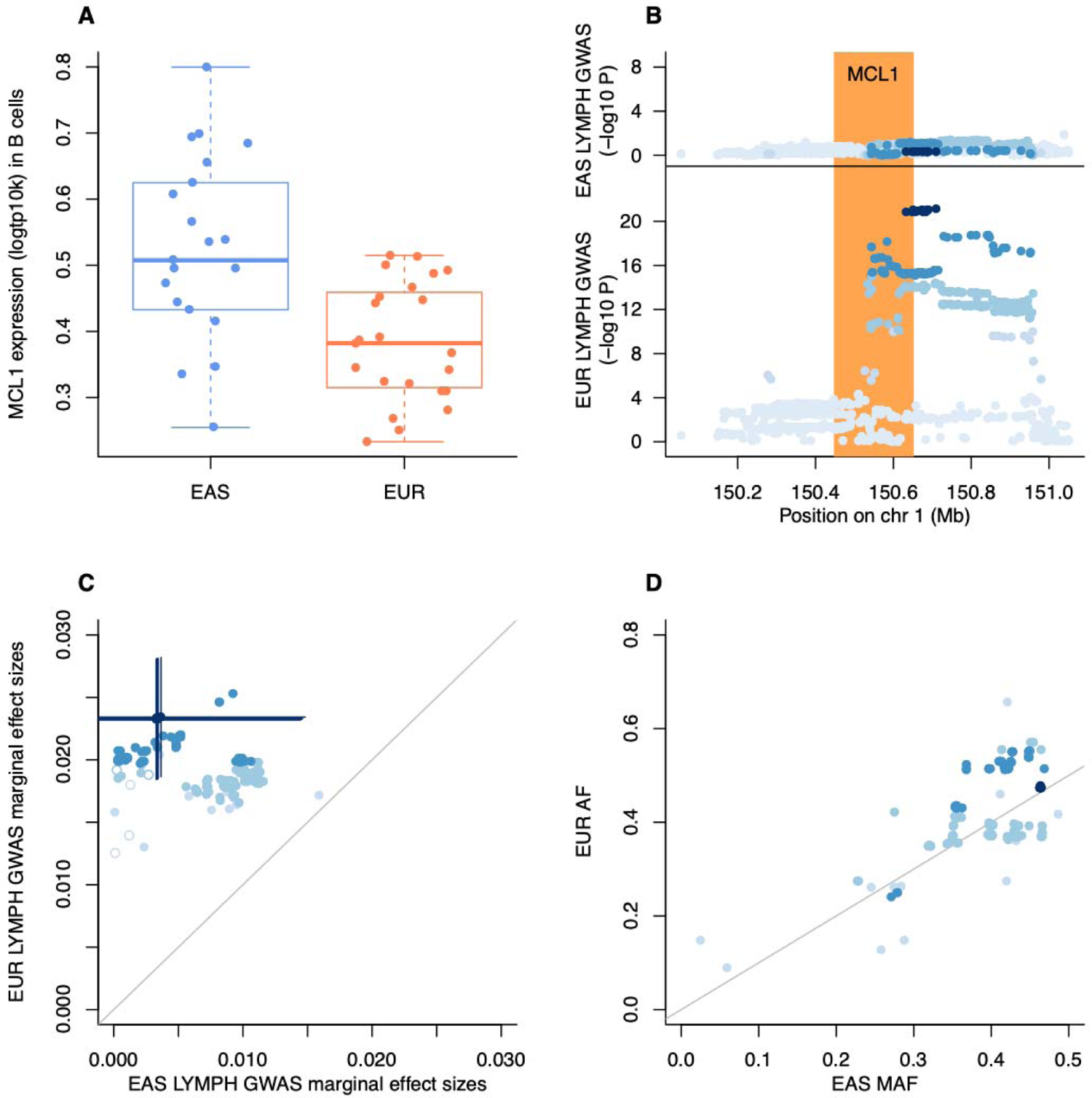
Discordant results between EAS and EUR Lymphocyte Count GWAS around the B-cell ancDE gene *MCL1*. **(A)** We report logtp10k for *MCL1* pseudo-bulk gene expression in B cells between ASI and EUR individuals. The median value of each expression is displayed as a band inside each box. Boxes denote values in the second and third quartiles. The length of each whisker is 1.5 times the interquartile range (defined as the height of each box). All dots represent observed values. **(B)** We report lymphocyte count (LYMPH) -log_10_ GWAS *P* values computed in EAS and EUR populations. The orange region represents 100kb windows on either side of *MCL1* gene body. **(C)** We report LYMPH marginal effect sizes computed in EAS and EUR populations. Marginal effects are plotted using absolute values. Blue lines represent 95% CI. **(D)** We report EAS MAF and EUR AF in 1000 Genomes. Color intensity in **(A,B,C)** represents GWAS -log_10_(*P*) differences between EAS and EUR.

*MCL1* is a gene that is essential to B cell development ^35–37^, which we only found significantly (FDR 5%) differentially expressed between EAS and EUR in B cells (*P* = 2 × 10^-5^; **Figure 6A** and **Supplementary Table 3**). While no *MCL1* eQTLs in B cells were reported in ref.^28^, we investigated *MCL1* eQTLs in blood in EAS (262 samples ^38^) and EUR (30,174 samples ^39^) datasets. We observed shared eQTLs (with similar allele frequency) across the datasets, but larger effect sizes in EAS (**Supplementary** Fig. 13), consistent with the higher expression of *MCL1* in B cells in EAS.

We observed significant associations in the LYMPH EUR GWAS (minimum *P* = 7 × 10^-22^ for rs6587520) around the *MCL1* gene, but not in the LYMPH EAS GWAS (*P* at rs6587520 in EAS = 0.52) (**Figure 6B**). We observed significant different marginal allele per-effect sizes at the EUR most significant loci (per-allele effect size of rs6587520 T allele = −0.004 ± 0.006 and −0.023 ± 0.002 in EAS and EUR, respectively; *P* = 1 × 10^-3^ for difference; **Figure 6C**), similar allele frequencies for the most associated variants in EUR (**Figure 6D**), and similar LD patterns with rs6587520 in both ancestries (**Supplementary** Fig. 14), demonstrating that these discordant effects were not driven by power issues due to different GWAS sample sizes (*N* = 62K in EAS ^40^ vs. *N* = 338K in EUR ^41^), and different allele frequencies and difference LD structure across the ancestries. One possible interpretation of these results would be that, because of the lower *MCL1* expression in European populations (inducing reduction of B cells ^35^), the variant rs6587520 will counterbalance this effect by increasing lymphocyte counts in European populations (as rs6587520 common allele increases lymphocyte counts, rs6587520 is expected to increase (on average) lymphocyte counts in European populations).

All together, these results suggest that GxE interactions might have led to ancestry-specific gene expression in B cells, ancestry-specific effect sizes of the gene eQTLs in blood, and ancestry-specific GWAS allele effect sizes in lymphocyte count around this gene.

## Discussion

Here, we characterized ancestry-specific gene regulation architectures at the cell-type level and investigated its overlap with ancestry-specific disease architectures. We analyzed scRNA-seq data in PBMCs from 44 individuals of EAS or EUR ancestry, and observed that ancDE genes tend to be differentially expressed in a single cell type, were enriched in genes involved in immune response to the environment, and that at least of third of ancDE genes could be due to allele frequency differences of their eQTLs (although it is likely that a large fraction of these eQTLs have allele frequency differences due to adaptation to new environment). Then, by leveraging ancestry-matched GWAS of 31 diseases and complex traits, we determined that squared multi-ancestry genetic correlation enrichment is λ^2^ = 0.69 ± 0.04 for SNPs surrounding ancDE genes, representing the lowest correlation reported by S-LDXR; numbers were similar when stratifying genes with and without eQTLs, suggesting that even if genes were differentially expressed due to allele frequency differences of their eQTLs, they are likely enriched in ancestry-specific effect sizes. We observed that these depletions were driven by ancDE genes from B cells (λ^2^ = 0.35 ± 0.06) and conventional dendritic cells (λ^2^ = 0.36 ± 0.10). Finally, we illustrated how GxE interactions may have led to differential expression of the ancDE gene *MCL1* in B cells, different *MCL1* eQTL effect sizes in blood, and different allele effect sizes around *MCL1* in LYMPH EAS and EUR GWAS.

To validate that cell-type-specificity of ancDE genes were not driven by low single-cell sample size and that our S-LDXR results were not driven by different allele frequency and LD structure across ancestries, we also extended our approach to sex-specific regulatory and complex trait architectures and observed similar patterns. Specifically, by detecting sexDE genes in a larger single-cell dataset ^28^ (1,175,543 cells from 982 donors), we showed that sexDE genes were also cell-type specific (**Supplementary** Fig. 4 and **Supplementary Table 6**), and that nearly a quarter have at least one independent eQTLs (**Supplementary** Fig. 8), demonstrating that genes with eQTLs can be differentially expressed even without differences in allele frequencies. Then, by extending S-LDXR to sex-specific effect sizes in sex-specific GWASs (in theory not subject to allele frequency and LD differences across investigated populations), we observed a significant depletion of squared sex genetic correlation within 17 independent male and female GWASs ^34^ (λ^2^ = 0.91 ± 0.02, *P* < 2 × 10^-7^) (**Supplementary Table 12**), confirming the impact of GxE interactions within GWAS effect sizes. In supplementary analyses, we also assessed discordant effects of sex-stratified GWAS within functional annotations from the baseline-LD model ^32^ and showed similar enrichment of squared multi-ancestry and sex genetic correlations across annotations (**Supplementary** Fig. 15).

Our findings have several implications for downstream analyses. First, they provide a partial source of explanation for the non-transportability of polygenic risk scores across populations ^6^. Accounting for ancDE genes in relevant cell types could help to downweigh variant effects when computing polygenic risk scores. Second, our results highlight the benefits of generating single-cell datasets (rather than functional dataset in bulk tissues) in non-European populations, as ancestry-specific regulation tends to be cell-type-specific. Finally, we proposed a framework leveraging single-cell and GWAS datasets that could be extended to analyze the impact of any environment interactions into complex traits, as performed here by considering ancestry and sex as environments.

We note several limitations of our work. First, although our dataset was (to our knowledge) the largest multi-ancestry scRNA-seq dataset publicly available, it contains only 44 individuals, which limited us to detect a hundred of FDR significant ancDE genes in the most abundant cell types. However, extremely significant S-LDXR results obtained in the top 100 ancDE genes in ncM (one of the lowest abundant cell types with 5,149 cells) suggest that the gene ranking of our analyses is robust, even if many of these genes are not significant at the FDR level. Low sample size also prevented us from performing eQTL analyses and to directly quantify ancestry-specific eQTL effect sizes at the cell-type level. In our application to the *MCL1* gene, we notably had to investigate blood eQTLs measured in non-homogenous EAS and EUR datasets, which might have impacted our conclusion on ancestry-specific eQTL effect sizes (although the observed effect was consistent with the higher expression of *MCL1* in B cells in EAS). Second, our analyses were restricted to populations of only two ancestries, which are the only ones with both large functional and GWAS datasets available. Ongoing efforts to generate both functional and GWAS datasets in diverse populations would help to replicate our results. Third, our analyses were restricted to gene expression, and did not investigate the impact of ancestry-specific regulatory elements (such as enhancers), whereas chromatin signals have been observed to be more ancestry-specific than gene expression ^24^. We anticipate that generating diverse functional datasets (such as single-cell ATAC-seq or single-cell multiome) in diverse ancestries will help to investigate ancestry-specific regulation at a finer scale. Fourth, our analyses were restricted to 7 main PBMC cell types, limiting the characterization of rarer cell types, as well as the characterization of the cellular composition of each main cell type. Despite these limitations, our results convincingly demonstrate that ancestry-specific effect sizes are enriched in genes with ancestry-specific regulation and demonstrate the urge to generate large single-cell and GWAS datasets in diverse populations to improve our understanding on the effect of genetic variants on human diseases.

## Data Availability

All data produced are available online at https://zenodo.org/records/10011016.

## Acknowledgment

We thank N. Mancuso, A. de Smith, A.L. Price, H. Shi, N. Zaitlen, C.J. Ye, R. Perez and G. Gordon for helpful discussion. S.G. and J. W. are funded by NIH grant R35 GM147789.

## Data and code availability

The single-cell dataset used in this study, differential gene expression results, S-LDXR files, summary statistics in EAS and EUR, and code to replicate our analyses are available at https://zenodo.org/records/10011016. Genotype data from ref. ^27^ is available in dbGap (dbGaP Study Accession: phs002812.v1.p1). S-LDXR is available at https://huwenboshi.github.io/s-ldxr/.

## Methods

### Single-cell RNA-seq data in peripheral blood mononuclear cells

We used processed scRNA-seq data in PBMCs from ref. ^27^, containing 256 individuals of East Asian (EAS) and European (EUR) ancestries. After restricting to 98 controls individuals and removing 158 systemic lupus erythematosus cases to have a homogeneous population, removing 50 individuals from the ImmVar study (which had only European individuals), 2 outliers in a principal component analysis (PCA) (**Supplementary** Fig. 16), and two males in the remaining dataset, we obtained a dataset of 21 EAS female controls and 23 EUR female controls generated in similar batches.

We restricted our analyses to the 7 most abundant cell types (between 18,203 and 380,477 cells per cell type before quality control), and removed cells labeled as PB (1,411 cells), Progen (807 cells), Prolif (8,265 cells), and pDC (5,233 cells). After removing cells with more than 20% of their reads in 13 mitochondrial (MT) genes, and cells with less than 500 reads or more than 10,000 reads, we obtained a total of 172,385 cells across the 7 cell types (**Supplementary Table 1**).

For supplementary analyses, we used the single-cell RNA-seq in 982 Europeans (565 females and 416 males) in PBMCs from ref. ^28^. We defined B cells as the ones labeled as “naive B cell”, “memory B cell” and “transitional stage B cell”, NK cells as the ones labeled as “natural killer cell”, T4 cells as the ones labeled as “central memory CD4-positive, alpha-beta T cell”, “naive thymus-derived CD4-positive, alpha-beta T cell”, “effector memory CD4-positive, alpha-beta T cell”, “CD4-positive, alpha-beta cytotoxic T cell”, “CD4-positive, alpha-beta T cell”, T8 cells as the ones labeled as “effector memory CD8-positive, alpha-beta T cell”, “naive thymus-derived CD8-positive, alpha-beta T cell”, “central memory CD8-positive, alpha-beta T cell”, “CD8-positive, alpha-beta T cell”, cM cells as the ones labeled as “CD14-positive monocyte” and ncM as the ones labeled as “CD14-low, CD16-positive monocyte”. We applied similar quality control than described above and obtained a total of 1,175,543 cells within the 7 cell types.

### Genes differentially expressed across ancestries

Within each cell type, we tested if each gene was differentially expressed between EAS and EUR individuals by using a Poisson linear mixed-effects model with the number of reads as the outcome variable, the donor as a random effect, and ancestry, age, batch, 5 first principal components of a principal component analysis (PCA) computed at the cell-type level on the 2,000 most variable genes, log of total number of reads per cell, proportion of genes expressed in a single cell (CDR: cellular detection rate) and fraction of reads in MT genes as fixed effect covariates. We restricted our analyses to 19,995 genes ^42^, and for each cell type we restricted our analyses to genes with at least 50 reads across all controls (48% of the genes on average across the cell types). For main analyses, we defined genes with the top 100 smallest *P* values for the ancestry covariate as ancDE genes. We computed FDR *P* values using the Benjamini-Hochberg correction ^43^ implemented in the R p.adjust function. We performed a similar approach to compute genes differentially expressed between females and males (sexDE genes) using data from ref. ^28^. We note that using a Poisson linear mixed-effects model with principal components computed at the cell-type level should have limited the impact of cell type heterogeneity on ancDE and sexDE gene results (as reported in refs. ^18,44^).

### Estimating enrichment of stratified squared multi-ancestry genetic correlation using S-LDXR

S-LDXR ^7^ is a method to estimate enrichment of stratified squared multi-ancestry genetic correlation across functional categories of SNPs using GWAS summary statistics and ancestry-matched linkage disequilibrium (LD) reference panels. S-LDXR models per-allele effect sizes (accounting for differences in allele frequency differences between populations) of SNP *j* in two populations (labeled as β_1j_ and β_2j_) with variance and covariance,

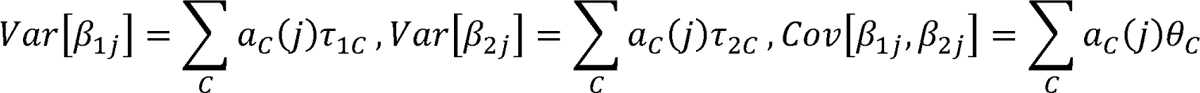

where *a_C_(j)* is the value of SNP *j* for annotation *C, τ_1C_* and τ_2C_ are the net contribution of annotation *C* to the variance of β_1j_ and β_2j_, respectively, and θ_C_ is the net contribution of annotation *C* to the covariance of β_1j_ and β_2j_.

S-LDXR estimates the stratified squared multi-ancestry genetic correlation, which is defined as

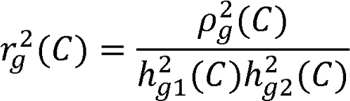

where *h^2^_g1_* and *h^2^_g2_* are heritabilities in each population, and is the multi-ancestry genetic covariance of each binary annotation *C*:

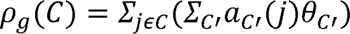

where *a_c’(j)_* and θ_c_ are annotations and coefficients for all annotations included in the analysis, respectively.

Then S-LDXR estimate the enrichment of squared multi-ancestry genetic correlation, which is defined as

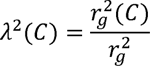

where *r^2^_g_* is the genome-wide squared multi-ancestry genetic correlation.

We applied S-LDXR using recommended settings ^45^, reference files (i.e., 481 East Asian and 489 European samples in the 1000 Genomes Project ^30^), and a background set of functional annotations (i.e., the baseline-LD-X model, a set of 62 functional SNP-annotations known to impact per-allele effect sizes). We applied S-LDXR on 31 diseases and complex traits^7^ (average N = 90K and 267K in EAS and EUR, respectively); most of the results were meta-analyzed across 20 approximately independent traits, including 10 approximately independent blood and immune-related traits (**Supplementary Table 2**). Reported *P* values for heritability enrichments were two-sided (i.e., testing if heritability enrichment is different from 1); reported *P* values for and were one-sided (i.e., testing if and are lower than 1 and 0, respectively). Our analyses included SNP-annotations related to gene sets, which were constructed by adding 100-kb windows on either side of the transcribed region of each gene in the set ^7,46^. All analyses included a SNP-annotation for the 19,995 genes, and seven SNP-annotations representing the set of genes expressed in each cell type (i.e., genes with at least 50 reads across all controls).

### Gene ontology (GO) enrichment analysis

We performed GO enrichment analysis of ancDE genes using R package *goseq* ^29^. We restricted analyzed pathways to the ones containing between 10 and 1,000 genes. We defined the reference set of genes as the genes with at least 50 reads across all samples within investigated cell types. We computed FDR *P* values using the Benjamini and Hochberg correction ^43^ implemented in the R p.adjust function.

### Cell-type-specific eQTL analyses

To determine whether the cell-type-specific ancDE genes were driven by allele frequency differences, we leveraged independent cell-type-specific eQTLs from ref. ^28^. We defined B cells eQTLs by merging eQTLs from cell types labeled as “B IN” and “B Mem”, T4 eQTLs by merging eQTLs from cell types labeled as “CD4 ET”, “CD4 NC” and “CD4 SOX4”, T8 eQTLs by merging eQTLs from cell types labeled as “CD8 ET”, “CD8 NC” and “CD8 S100B”. We defined cDC, cM, nCM and NK eQTLs by considering eQTLs from cell types labeled as “DC”, “Mono C”, “Mono NC” and “NK R”, respectively. We restricted all analyses to variants with a MAF > 5% in EUR.

We compared the EAS and EUR allele frequency of these eQTLs using 481 EAS and 489 EUR individuals from 1000 Genomes ^30,47^. Fixation index (*F_st_*) across EAS and EUR populations were computed using the formula ^48^

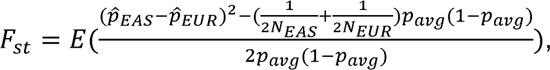

where *p_EAS_*and *p_EUR_* are the allele frequencies estimated in the EAS and EUR populations, respectively, *N_EAS_* and *N_EUR_* are the sample size of these populations, and *p_avg_* is defined as *(p_EAS_ + p_EUR_)/2*

To test if the 329 ancDE genes with EUR independent eQTLs were still significantly differentially expressed after conditioning to the genotypes of these eQTLs, we extracted those SNPs in the genetic data of ref. ^27^ (genotypes of eQTLs were available for 273 out of 329 genes) and replicated our differentially expression analyses for ancDE genes while correcting for the genotypes of the eQTLs.

### Extending S-LDXR to estimate enrichment of stratified squared sex genetic correlation

We extended S-LDXR to estimate enrichment of stratified squared sex genetic correlation using GWAS summary statistics computed in males and females of the same ancestry, and a corresponding LD reference panel. Here, we applied S-LDXR using recommended settings ^45^, and the EUR reference file and the baseline-LD model version 2.2 used by S-LDSC ^32^. We downloaded the male and female GWASs previously identified with sex genetic correlation significantly less than 1 (ref. ^34^), and defined a set of 17 independent traits with genetic correlation ^49^ < 0.1. We observed consistent squared sex genetic correlation and multi-ancestry genetic correlation among annotations of the baseline-LD and baseline-LD-X models (**Supplementary** Fig. 15).

